# Genome-wide analysis of 64 male reproductive phenotypes reveals novel loci and shared biological pathways

**DOI:** 10.64898/2025.12.20.25342734

**Authors:** Jéssica Figuerêdo, Toomas Haller, Estonian Biobank Research Team, Health Informatics Research Team, Margus Punab, Triin Laisk, Reedik Mägi

## Abstract

Male reproductive disorders affect millions of men worldwide, yet their genetic basis remains poorly understood. Conditions such as infertility, erectile dysfunction, prostate disorders and hormonal abnormalities are common, interconnected and often diagnosed late, underscoring the need for broader biological insight. Using genetic data from over 530,000 men across the Estonian Biobank, FinnGen, and UK Biobank, we analysed 64 male reproductive phenotypes. We identified 143 lead variants associated with these conditions, including 47 not previously linked to male reproductive traits, and detected the first genome-wide signals for 3 understudied disorders. Twelve lead variants showed at least 1.5-fold enrichment in the Finnish and Estonian populations, indicating population-specific effects and utility of isolated cohorts for variant discovery. We also highlighted 328 candidate genes and mapped shared biological pathways across prostate, testicular, penile and hormonal traits. These findings expand knowledge of male reproductive biology and offer entry points for improved diagnosis and risk prediction.

## INTRODUCTION

Male reproductive health is a key component of overall well-being, yet it remains underrepresented in genomic research on complex traits. This gap largely reflects challenges in defining and capturing many male reproductive phenotypes at population scale, rather than a lack of clinical relevance (1). Conditions such as infertility, sexual dysfunction, prostate diseases, and hormonal imbalances, are common and often interconnected, affecting men across lifespan and contributing to substantial long-term health, psychological, and quality-of-life burden (2). Despite growing awareness of their clinical and societal importance, the genetic architecture of many male-specific reproductive conditions remains poorly understood.

The expansion of large population-based biobanks, including the Estonian Biobank (EstBB), Finnish biobank research project (FinnGen), and UK Biobank (UKBB) have become essential for mapping the genetic basis of complex traits. Their scale, combined with longitudinal follow-up and linkage to electronic health records (EHRs), enables well-powered genome-wide association studies (GWAS) across a broad range of phenotypes (3–5). The use of standardized coding systems, such as the International Classification of Diseases (ICD) for diagnoses and Logical Observation Identifiers Names and Codes (LOINC) for laboratory measurements, further supports consistent and reproducible phenotype definitions across cohorts (6,7). Together, these structures make it possible to harmonize analyses across studies and to perform meta-analyses that improve robustness and generalizability. Recent work using these resources has uncovered shared and population-specific variants for a wide range of traits, including metabolic, cardiovascular, and reproductive conditions (8–11). In particular, genetically isolated populations such as those of Finland and Estonia, offer increased power to detect risk variants that may be undetected in more heterogeneous cohorts (4).

Despite these advances, prior GWAS of male reproductive health traits have concentrated on a limited number of traits. Testosterone levels, erectile dysfunction, and prostate cancer account for much of the existing literature, with multiple genome-wide associations reported for these phenotypes (9,11,12). By contrast, many genitourinary and endocrine conditions affecting male reproductive function have received less attention in large-scale genetic studies. As a result, sex-specific disease mechanisms and pleiotropic effects across reproductive phenotypes remain poorly resolved, reflecting limited trait coverage and the lack of well-powered, male-focused datasets (1).

To address these limitations, here we present the largest and most comprehensive GWAS and meta-analysis of male reproductive phenotypes to date, leveraging data from EstBB, FinnGen, and UKBB (Figure 1). Across all analyses, the combined meta-analysis sample size reached up to 594,127 individuals, depending on phenotype availability (Supplementary table 2). 64 male reproductive phenotypes were examined, representing the full set of traits available across cohorts that met predefined inclusion and power criteria. These phenotypes encompass major domains of male reproductive health, including neoplastic, endocrine, structural, inflammatory, and functional traits, and were defined using curated ICD-10 and LOINC codes. By integrating variant-level association, colocalization, gene prioritisation, pathway enrichment, and genetic correlation analyses, we identified 143 genome-wide significant variants, including 47 novel trait-specific associations, and uncovered the first genome-wide signals for previously unexplored conditions namely abnormal spermatozoa, phimosis, and testicular dysfunction, while prioritising 328 candidate causal genes that highlight shared biological mechanisms across prostate, testicular, and penile traits.

**Figure 1.**
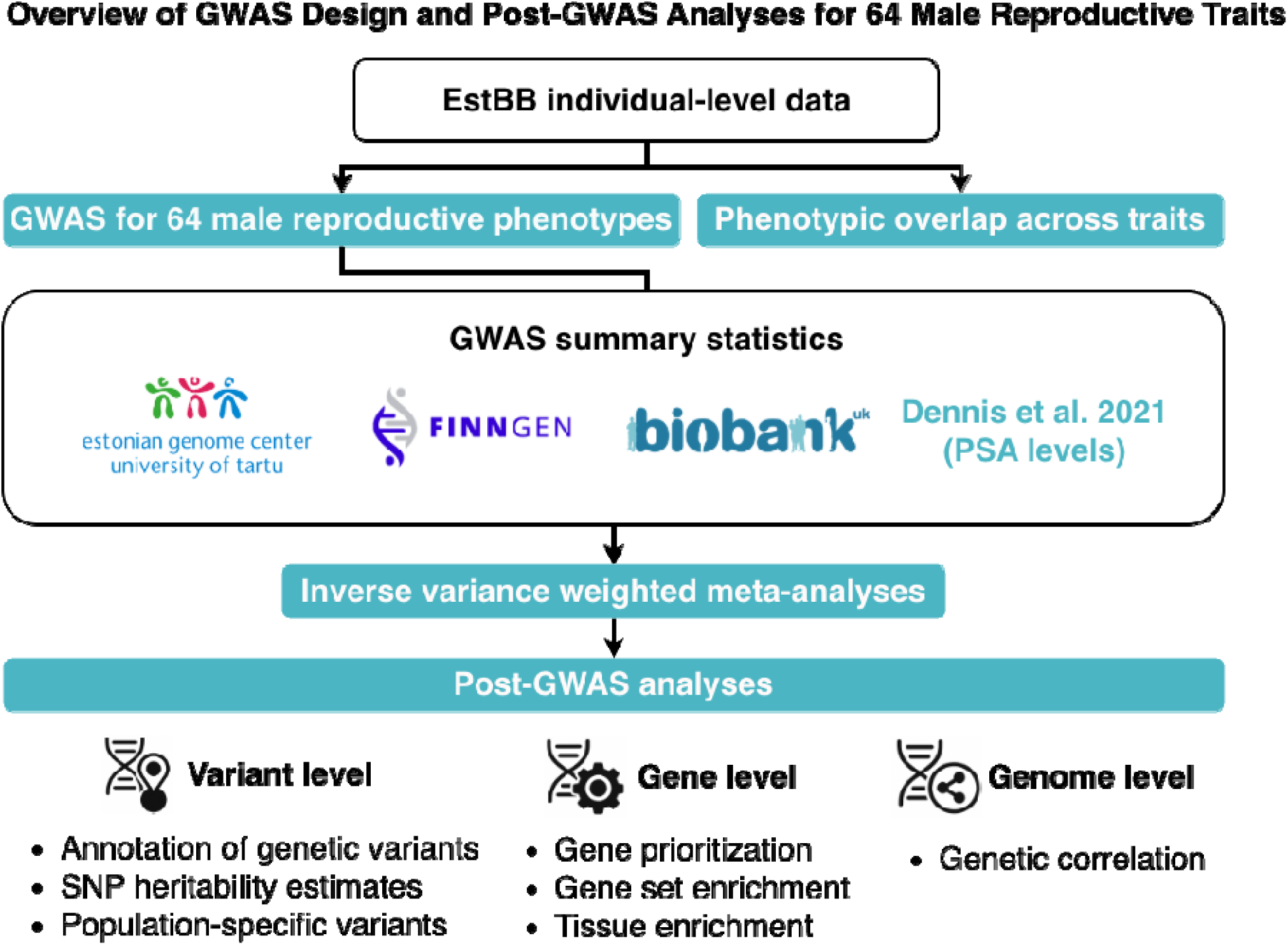
Flowchart of the study design and analysis workflow. The study comprised three main steps: (1) genome-wide association studies (GWAS) for 64 male reproductive phenotypes using individual-level data from the Estonian biobank (EstBB); (2) phenotypic overlap across traits using individual-level data from the EstBB; (3) GWAS meta-analyses incorporating summary statistics from EstBB, FinnGen, UK Biobank and/or Dennis et al. (2021); and (4) post-GWAS analyses performed at three levels: variant, gene and genome. PSA= prostate-specific antigen.

## METHODS

### Estonian Biobank cohort, phenotype definition and GWAS analysis

EstBB is a population-based cohort of approximately 212,000 participants (∼20% of the Estonian adult population) with extensive phenotypic and health-related information collected for each individual (3,13). At recruitment, participants provide broad consent allowing longitudinal linkage to national health registries and EHRs, including laboratory measurements. EstBB is regularly linked to the National Health Insurance Fund database (from 2004 onwards), as well as other national registries, including population, mortality, cancer, and prescription databases. For each participant, diagnostic information is available using ICD-10 codes (3).

Genotyping of DNA samples from the EstBB was done at the Core Genotyping Laboratory of the Institute of Genomics, University of Tartu using the Illumina Global Screening Array v3.0_EST. Altogether 295,810 samples were genotyped and then PLINK format files were created using Illumina GenomeStudio v2.0.4 (14). During the quality control, individuals were excluded if they had a call-rate < 95%, showed discordance between reported sex and genetically inferred sex based on X-chromosome heterozygosity, or were outliers in autosomal heterozygosity test. Variants were filtered by call-rate < 95% and Hardy-Weinberg equilibrium P < 1 × 10^-4^ (autosomal variants only) (14). Genotyped variants positions were in build 37 and were lifted over to build 38 using Picard (https://broadinstitute.github.io/picard/). Before imputation, variants with minimum allele frequency (MAF) < 1% and indels were removed. Phasing and imputation were done using Beagle v5.4 (beagle v.22Jul22.46e) and default settings (15). An Estonian population-specific imputation panel based on 2,695 whole-genome-sequenced samples was used as the reference, together with standard Beagle hg38 recombination maps (14). Based on principal components (PCs) analysis, samples that were not of European ancestry were excluded. Duplicate samples and monozygotic twins were identified using Kinship-based Inference for GWAS (KING) 2.2.7 (16), and one individual from each pair of duplicates was excluded.

Briefly, we used ICD-10 codes to identify individuals for male health phenotypes. Cases and controls were selected using ICD-10 codes for 52 male health phenotypes. Controls were individuals with no evidence of the analysed phenotypes, as defined in this study. Detailed information on the ICD codes/phenotypes used and cases and controls definitions and sample size are in the supplementary tables 2 and 3.

For the GWAS of male reproductive health-related laboratory parameters we selected 12 measurements of LOINC codes obtained from EHRs between the period of 2008-2020. More information on the codes and measurements used can be found in the supplementary table 3. During quality control, observations lying more than three standard deviations from the mean were excluded. The raw data extraction workflow can be found in the supplementary information (Text S3) of Kurvits et al. (2023). For individuals with multiple measurements, the most recent available measurement was used for GWAS analysis.

We conducted GWASs for the defined phenotypes (up to 67,744 men) using the REGENIE software (version 3.2) (18) adjusting for the first ten principal components, and year of birth.

### FinnGen cohort, phenotype definition and GWAS analysis

FinnGen research project is a public-private partnership which contains 392,000 Finnish individual’s genome information combined with digital health record data (4). The data release R8 (https://r8.finngen.fi/) was used for most of the analyses presented here. The data release R7 (https://r7.finngen.fi/) was used for two phenotypes (“Abnormal spermatozoa” and “Testicular dysfunction”), because they were not present in data release R8. Individuals in the FinnGen study were genotyped using Illumina and Affymetrix arrays (Illumina Inc., San Diego, CA, USA; Thermo Fisher Scientific, Santa Clara, CA, USA). Genotype imputation was performed with the Finnish population-specific SISu v3 reference panel, based on 3,775 whole-genome sequences. Disease endpoints were obtained from nation-wide health registries. GWAS analyses were conducted using a modified version of REGENIE v2.0.2, which includes dosage-based allele frequency calculation in cases and controls. The GWAS models were adjusted for age, 10 PCs and genotyping batch as covariates (4).

A total of 22 endpoints were available for the selected phenotypes and their GWAS summary statistics were used for the analyses. See Supplementary Table 4 for phenotype definitions.

### UK Biobank cohort, phenotype definition and GWAS analysis

UKBB is a large population-based study that has genome and phenotype data for more than 500,000 individuals (5,19). For the meta-analysis, GWAS summary statistics were obtained via the PheWeb portal (https://pheweb.org/UKB-TOPMed/phenotypes) which contains EHRs-derived ICD codes from the White British participants of the UKBB. Genotype data had been imputed using Trans-Omics for Precision Medicine (TOPMed) and GWAS analyses were conducted using SAIGE adjusting for genetic relatedness, sex, year of birth and the first 4 PCs (20,21). A total of 19 phenotypes with available GWAS summary statistics were used in this study. See Supplementary Table 4 for phenotype definitions.

### GWAS meta-analysis

GWAS meta-analyses were conducted for 27 phenotypes according to the availability of the phenotypes in the EstBB, FinnGen, and UKBB cohorts (Supplementary table 2, Figure 1). The “Prostate-specific antigen levels” (PSA levels) phenotype differed from other analyses in that it combined EstBB data with GWAS summary statistics from Dennis et al. (22), including 7,837 individuals of European ancestry (GWAS Catalog: GCST90012723). Phenotype and cohort specific sample sizes are reported in Supplementary Table 2.

Meta-analyses were done using the inverse of variance based fixed effect method implemented in Genome-Wide Association Meta-Analysis (GWAMA) (23). Genome-wide statistical significance in each analysis was defined as P < 5 × 10^-8^ and only variants that were present in at least two cohorts across all meta-analyses and passed the MAF filter of 1% were used in the follow-up analysis. Additionally, we report the number of genome-wide significant variants that met a stricter study-wide significance threshold of P < 5 × 10^-8^/64 = 7.81 × 10^-10^, accounting for the total number of phenotypes analysed. Heterogeneity for the variants was evaluated using *I^2^* index by Higgins et al. (24) implemented in GWAMA.

### Annotation of GWAS signals

Annotation of GWAS signals was done using the Functional Mapping and Annotation of Genome-Wide Association Studies (FUMA) platform (v1.3.6a) (25). FUMA defines genomic risk loci from independent significant single nucleotide polymorphisms (SNPs) by merging linkage disequilibrium (LD) blocks, gene mapping and annotates (using ANNOVAR) all input variants.

Signals with lead variants rs184810529 and rs4907778 from “Malignant neoplasm of prostate” GWAS Meta-analysis were excluded since they were present in only one cohort. Variants located inside the Major Histocompatibility complex (MHC) region were not annotated due to the complex LD structure. Thus, the genomic locus with lead variant rs12662632 from “Redundant prepuce, phimosis and paraphimosis” was excluded from the follow-up analysis, along with the three signals (lead variants rs2523570, rs28371333, and rs9277621) in the human leukocyte antigen (HLA) region detected in the GWAS meta-analysis for the phenotype “Disorders of penis”. “Urethritis and urethral syndrome” showed two genome-wide significant signals, but they were not included in the annotation, because: (1) the signals were singletons (no other SNPs have r^2^ > 0.6); (2) there was no supporting evidence available from the biological perspective for the variants and candidate genes; (3) the analysis was done using only EstBB data and we lacked a suitable replication dataset.

Novel loci were defined as those showing no prior association with the corresponding phenotypes. Loci were delineated by the lead variant, variants in LD (*r*^2^ > 0.6; FUMA-based), and SNPs within a 1Mb window around the lead variant. Evidence for prior associations was assessed using GWAS Catalog lookups implemented in FUMA, complemented by direct searches of the GWAS Catalog website and relevant published literature.

### Gene prioritisation

To identify candidate causal genes at each genomic risk locus, we performed gene prioritisation based on four criteria: 1) proximity to the lead variant; 2) presence of missense variants within the gene, or in high LD (r^2^ > 0.6) with it; 3) significant colocalization (posterior probability ≥ 0.8) between GWAS signal and gene expression (expression quantitative trait locus; eQTL) in relevant tissues; and 4) evidence of reproductive-related phenotypes in mouse models or involvement in human monogenic diseases.

For the Nearest gene parameter (proximity to the lead variant), protein-coding genes were identified using both FUMA and the Open Target Genetics platform (26,27). When results differed between these tools, priority was given to genes previously associated with reproductive health traits in the GWAS Catalog or supported by additional evidence from our prioritisation framework.

Candidate genes were ranked based on the number and relative strength of supporting criteria. Genes supported by multiple lines of evidence were prioritised over those supported by a single criterion. When only one criterion was available for a given locus, candidate genes were selected according to a predefined hierarchy of evidence strength, ordered were weighted as follows: 1) association with reproductive health phenotypes in mouse model; 2) eQTL-GWAS colocalization; 3) GWAS signal tagging a missense variant within the gene; and 4) proximity to GWAS signal. This hierarchy reflects decreasing evidence specificity for causal involvement.

#### a. Correlation with mouse phenotypes

The Mouse Genome Database (MGD; https://www.informatics.jax.org/) was used to assess whether candidate genes identified in this study were associated with reproductive health-related phenotypes in mouse models or with monogenic human diseases affecting female or male reproductive traits. MGD integrates mouse phenotypes annotations with human gene-disease relationships from the National Center for Biotechnology Information (NCBI) and Online Mendelian Inheritance in Man (OMIM), as well as human disease-to-phenotype from the Human Phenotype Ontology (HPO) (28).

#### b. Colocalization

Colocalization analyses between GWAS summary statistics and the eQTL Catalog were performed using HyprColoc (29,30). Genotype-Tissue Expression (GTEx) v8 datasets for blood, prostate and testis tissues were used (dataset IDs: QTD000356, QTD000306, and QTD000336, respectively) (31). Lead SNPs were defined using a P threshold of 5 × 10^-8^, and colocalization was tested within a 1Mb window around each lead variant.

### SNP heritability

To calculate the SNP-based heritability (h^2^SNP) for the phenotypes we used LD Score regression (LDSC) v1.0.1 regression implemented in Complex Trait Genetics Virtual Lab (0.4-beta) (32,33). For this analysis, we used only the phenotypes that presented effective sample size [Neff = 4/((1/N total cases) + (1/N total controls))] greater than 5,000 in total (N=14). Observed-scale heritability estimates and their standard errors were converted to the liability scale using the formula described in Lee et al. (34), implemented in R v4.2.2. Population prevalence was assumed to equal the sample prevalence (i.e., the proportion of cases in the meta-analysis). Calculated sample prevalences are shown in the Supplementary Table 8.

### Genetic correlation analysis

We used LD Score regression implemented in the Complex Trait Genetics Virtual Lab (CTG-VL, version 0.4-beta) (32,33,35) to estimate genetic correlations. To account for multiple testing, we applied the Benjamini–Hochberg procedure using p.adjust function in R v4.2.3 to control the false discovery rate (FDR), considering associations with FDR-adjusted P < 0.05 as statistically significant.

Genetic correlations were performed for all male phenotypes with an effective sample size greater than 5,000, using two sources of GWAS summary statistics: (1) reproductive health-related hormone level GWAS datasets from the GWAS Catalog and published studies by Kachuri et al. (2023), Pott et al. (2019), Ruth et al. (2021), and Schumacher et al. (2018) (Supplementary table 16); and (2) 1,436 traits from the CTG-VL database, including eight lifestyle factors: smoking status (current, former and never), body mass index (BMI), educational attainment, and alcohol drinker status (current, former, and never). These traits were included to assess shared genetic architecture and pleiotropy between male reproductive traits and health-related lifestyle factors, which are partly modifiable.

Several phenotypes were excluded from the analysis due to their nonspecific definitions and substantial overlap with more clearly defined phenotypes included in the study. These comprised “Failure of genital response”, “Other disorders of penis”, “Impotence of organic origin”, “Disorders of prostate, unspecified”, “Hydrocele and spermatocele”, and “Orchitis, epididymitis and epididymo-orchitis without abscess”

### Phenotype-level correlation analysis

To assess phenotype-level associations and evaluate the correspondence between genetic and phenotypic correlations, we examined case-level co-occurrence among binary clinical phenotypes included in the genetic correlation analysis (see above). Only individual-level data from the EstBB were used for this analysis. Case overlap was quantified using the Jaccard Index, calculated as c11 / (c10 + c01 + c11), where c11 is the number of individuals with both diagnoses, and c10 and c01 are the numbers of individuals with only one of the two diagnoses. Lifestyle (e.g., alcohol consumption, smoking status), behavioural (e.g., educational attainment), and quantitative (e.g. BMI) traits were excluded, as they are not defined as case-control variables. Results were visualized using the corrplot package in R (v0.92; https://github.com/taiyun/corrplot).

In addition, given prior evidence linking male infertility to cancer (36), we specifically evaluated the phenotypic overlap between “Male infertility” and malignant neoplasms. Cancer cases were defined using all ICD-10 C-codes from the malignant neoplasms category (C00-C97), with all remaining individuals defined as controls. Since the EstBB EHRs are available from 2004 onwards, the analysis was restricted to individuals aged 18-40 years at recruitment, corresponding to the reproductive age range. The association was tested using logistic regression, adjusting for age at recruitment and the first ten genetic principal components.

### Gene-set and tissue enrichment

To investigate the biological pathways and tissue-specific expression patterns underlying male reproductive health phenotypes, we conducted gene-set and tissue enrichment analyses using FUMA, which implements Multi-marker Analysis of GenoMic Annotation (MAGMA) v1.08 for gene-property and gene-set analyses (25,37). MAGMA gene-set analysis was performed to test for enrichment of associated genes in predefined biological processes and molecular functions from the Gene Ontology (GO) database (38). MAGMA tissue expression analysis was performed using gene expression data from 53 tissue types of GTEx v8 to test for preferential expression of phenotype-associated genes. All analyses were run using default parameters, and statistical significance was assessed after Bonferroni correction for the number of tested gene sets or tissues.

### Population-specific variants

To identify population-specific variants in Finnish and Estonian populations, allele frequencies of lead variants were compared with those in broader European populations (non-Finnish Europeans). Population-specific minor allele frequencies were obtained from the Open Targets Genetics portal (https://genetics.opentargets.org/). Allele frequency enrichment was defined as the ratio of the minor allele frequency in the Finnish or Estonian populations to that in non-Finnish Europeans. Variants showing more than 1.5-fold enrichment were considered potentially population-specific (Supplementary table 7).

## RESULTS

### GWAS of male reproductive health traits

A total of 11 phenotypes showed genome-wide significant signals, where 143 genome-wide significant lead variants were identified (Supplementary table 5), 102 of them passing a multiple-testing threshold (P < 7.81 × 10^-10^) accounting for the number of tested phenotypes. The mean genomic inflation factor across all traits was 1.01 (range from 0.94 to 1.13) in binary traits and 0.94 (0.89 to 1.02) in quantitative traits, indicating that the test statistic inflation was well controlled.

We assessed the heterogeneity of genetic effects across studies using the Cochran’s Q-test and the corresponding I² statistic for each lead SNP (Supplementary table 5). Overall, most genome-wide significant loci showed low heterogeneity, indicating consistent effects across cohorts (EstBB, FinnGen and UKBB). However, 17 loci exhibited high heterogeneity (I^2^ > 70%), possibly due differences in effect allele frequencies, phenotype definitions, or cohort-specific ascertainment and data collection procedures (Supplementary table 5; Supplementary figure 1).

Based on systematic comparison with previously reported GWAS findings, 47 trait-specific loci were classified as novel for the corresponding phenotypes. Among these, 26 variants have not been previously associated with any reproductive trait (Figure 2; Supplementary table 6). Three phenotypes were associated with significant genome-wide association signals for the first time: “Abnormal spermatozoa” (lead variant: rs17632542), “Redundant prepuce, phimosis and paraphimosis” (lead variant: rs11198372), and “Testicular dysfunction” (lead variant: rs1799941) (Table 1).

**Figure 2.**
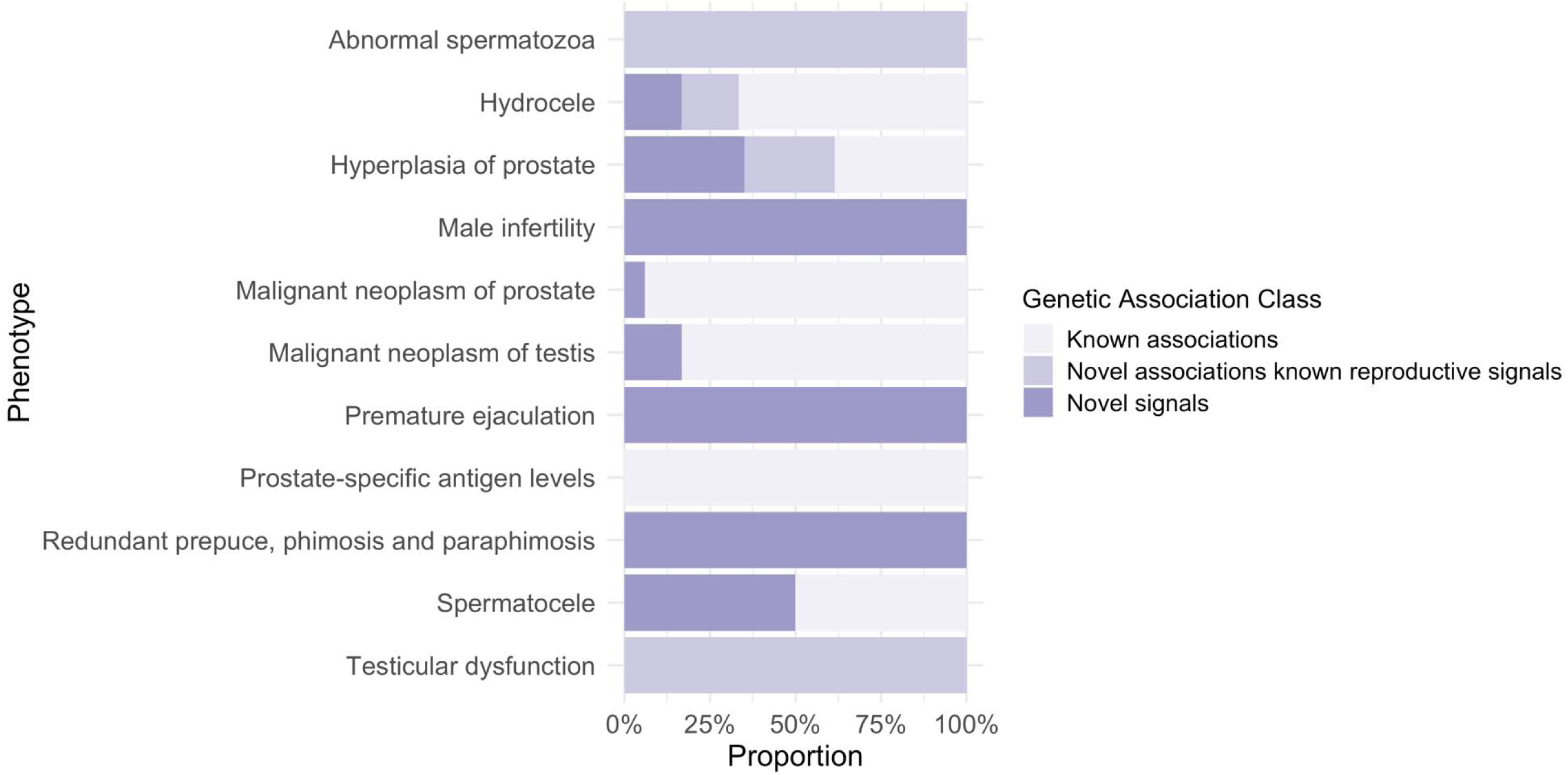
Overview of the known and novel genetic associations across male reproductive health phenotypes.

**Table 1.** A summary of first time reported GWAS and GWAS meta-analyses results for male reproductive health phenotypes.

| ABNORMAL SPERMATOZOA |  |  |  |  |  |  |  |  |
| --- | --- | --- | --- | --- | --- | --- | --- | --- |
| Chr (band) | POS (hg19) | Lead variant | EA/NEA | EstBB | EAF FinnGen | UKBB | OR (95% CI) | P-value |
| chr19(q13.33) | 51361757 | rs17632542 | T/C | 0.06 | 0.07 | 0.07 | 1.50 (1.32-1.71) | $8.83 \times 10^{-10}$ |
| REDUNDANT PREPUCE, PHIMOSIS AND PARAPHIMOSIS |  |  |  |  |  |  |  |  |
| Chr (band) | POS (hg19) | Lead variant | EA/NEA | EAF |  |  | OR (95% CI) | P-value |
|  |  |  |  | EstBB | FinnGen | UKBB |  |  |
| chr6(p21.32) | 32578383 | rs12662632 | G/C | 0.05 | 0.03 | 0.02 | 1.78 (1.66-1.92) | $6.13 \times 10^{-53}$ |
| chr10(q26.11) | 120108324 | rs11198372 | G/A | 0.59 | 0.63 | NA | 1.11 (1.07-1.15) | $7.74 \times 10^{-10}$ |
| TESTICULAR DYSFUNCTION |  |  |  |  |  |  |  |  |
| Chr (band) | POS (hg19) | Lead variant | EA/NEA | EAF |  |  | OR (95% CI) | P-value |
|  |  |  |  | EstBB | FinnGen | UKBB |  |  |
| chr17(p13.1) | 7533423 | rs1799941 | G/A | 0.29 | 0.27 | 0.26 | 1.31 (1.21-1.42) | $1.45 \times 10^{-10}$ |

**Abbreviations:** Chr, chromosome; POS, genomic position (hg19); EA, effect allele; NEA, non-effect allele; EAF, effect allele frequency; OR, odds ratio; CI, confidence interval.

### Gene prioritisation

A total of 699 genes showed at least one level of evidence of being associated with male phenotypes (Supplementary table 11). Notably, 8 genes prioritised for 4 phenotypes presented 3 levels of evidence: “Hyperplasia of prostate” - *CHEK2* (novel locus), *FOXF1* (novel locus), *POU5F1B*, *RSPO2* (novel locus), and *TFAP4* (novel locus); *“*Malignant neoplasm of prostate” - *AR* (novel locus), *LMTK2*, and *POU5F1B*; “Spermatocele” - *PAX8*; “Prostate-specific antigen levels” - *POU5F1B*.

All genes with three levels of evidence have been previously linked to reproductive traits and have well-characterized roles in reproduction. For example, *RSPO2* (*R Spondin 2 or roof plate-specific spondin-2*) regulates Wnt signaling and plays a key role in developmental processes and morphogenesis (39). Due to its function as an oncogenic driver, *RSPO2* has been widely implicated in various human carcinomas, including prostate cancer (12,39). Similarly, *CHEK2* (*checkpoint kinase 2*), a known tumour suppressor gene encoding a protein kinase activated in response to DNA damage, has been associated with several reproductive health traits, including androgen index free, breast cancer, female infertility, polycystic ovary syndrome, prostate carcinoma, PSA, testosterone levels, sex-hormone binding globulin, and uterine fibroids (9–12,40–43).

Based on our gene prioritisation approach, we identified 328 candidate causal genes across all the genomic risk loci (Supplementary table 11). Among these, four genes (*FGFR2*, *KLK3*, *POU5F1B*, and *TERT*) were prioritised for several prostate-related phenotypes, suggesting potential shared genetic mechanisms (Figure 3). *KLK3* and *POU5F1B* showed consistent links with prostate hyperplasia, prostate cancer, and PSA levels, in line with their established roles in prostate biology, while *TERT* was additionally associated with testicular malignancy, suggesting a broader involvement in male reproductive cancers. The recurrence of these genes across several phenotypes highlights their potential relevance to shared biological mechanisms underlying male reproductive function and disease.

**Figure 3.**
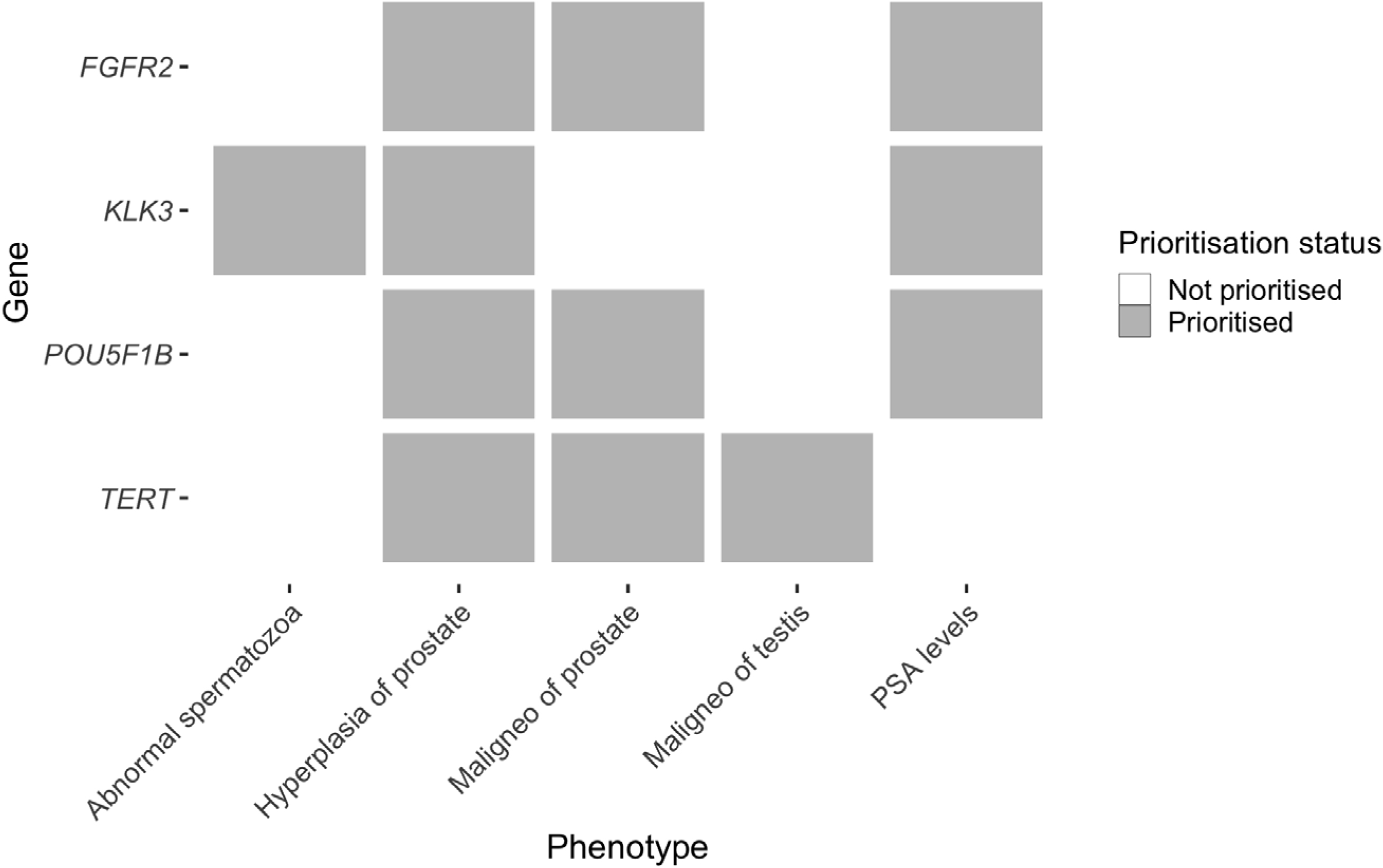
Recurrence of prioritised genes across prostate-related traits. Heatmap indicates whether a gene was prioritised as a candidate causal gene for each phenotype based on the multi-criteria framework described in Methods. Only genes associated with multiple prostate-related traits are shown. Maligneo= Malignant neoplasm; PSA= Prostate-specific antigen.

### Population-specific variants

Effect allele enrichment analyses in the Finnish and Estonian populations identified 12 variants with at least 1.5-fold enrichment compared to the broader European reference population, suggesting population-specific effects (Supplementary table 7). Among these, 4 variants were novel for the studied phenotypes, and 7 candidate genes are linked to phenotypes for the first time. Specifically, *MAF* and *WDR35* were newly associated with “Hyperplasia of prostate”, while *MYC*, *WASHC5*, *PNPO*, *LRRC46* and *ITGB3* were linked to “Malignant neoplasm of prostate”. These findings underscore the contribution of isolated populations in uncovering previously undetected disease loci.

For “Malignant neoplasm of prostate”, several lead variants exhibited population-specific allele enrichment. The strongest signal was a missense variant on chromosome X (rs200737258), which displayed an 8.6-fold higher enrichment in the Finnish population and a 3.2-fold higher frequency in Estonian population compared with non-Finnish Europeans. This variant represents a novel genome-wide signal mapped to the androgen receptor (AR) locus. Additional population-enriched variants included rs77482050 on chromosome 2, which showed 6.5-fold enrichment in Finns and 1.9-fold in Estonians, and rs191767420, associated with “Hyperplasia of prostate”, with 2.9-fold and 1.7-fold in Finnish and Estonian populations, respectively (Supplementary table 7).

### SNP-Heritability

Heritability from common SNPs was calculated for 27 phenotypes with an effective sample size > 5,000. Of these, 16 phenotypes showed statistically significant SNP-based heritability (P < 0.05 from chi-square test; Figure 4; Supplementary table 8), with liability-scale estimates ranging from 2% to 13%. Among the reproductive health traits, “Spermatocele” had the highest SNP-heritability estimate (13%) followed by “Malignant neoplasm of prostate” (12%). “Other disorders of prostate” (3%), “Inflammatory diseases of prostate” (2%), and “Other disorders of male genital organs” (2%) exhibited the lowest estimates, possibly reflecting a greater contribution of non-genetic factors to these phenotypes.

**Figure 4.**
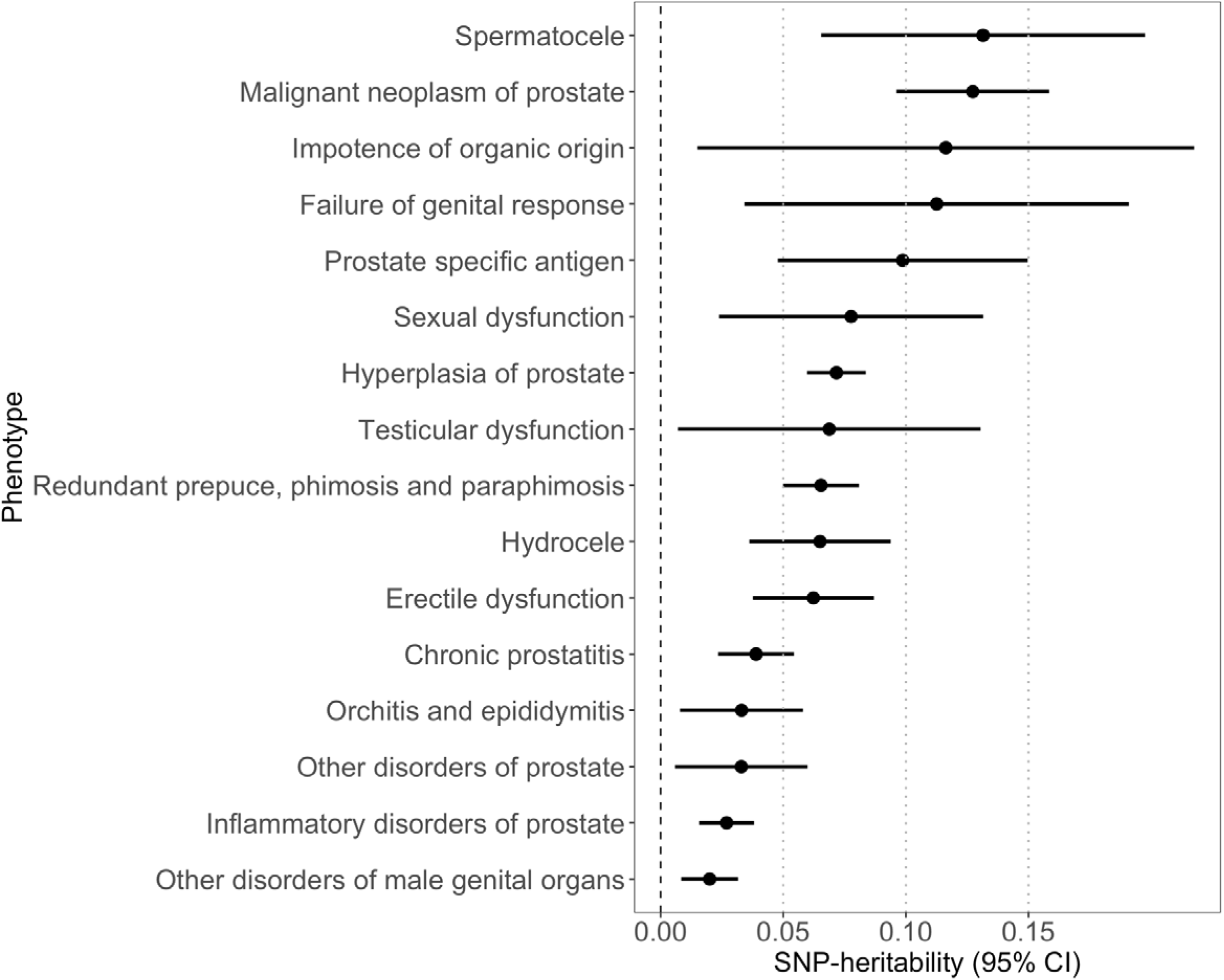
Statistically significant SNP heritability (P < 0.05, chi-square test) for male reproductive phenotypes with effective sample size > 5,000.

### Gene-set and tissue enrichment

MAGMA gene-set enrichment analysis across male reproductive health phenotypes identified several biological pathways enriched for associated genes (Supplementary tables 9 and 10). “Hyperplasia of prostate” had the largest number of significantly enriched gene sets, including pathways related to transcriptional regulation (e.g. “DNA-binding transcription factor activity” (P_bon_ = 3.82 × 10^-4^), transcriptional repressor complexes, cell cycle pathways, and fibroblast growth factor receptor signalling. These enrichments indicate that genes involved in transcriptional regulation and cell cycle-related process are overrepresented among loci associated with prostate hyperplasia. Similarly, “Malignant neoplasm of prostate” and “Phimosis” showed enrichment for gene sets involved in transmembrane transport and immune-related molecular functions, respectively.

To further explore the tissue-specific expression of genes associated with male reproductive health, we performed MAGMA tissue expression analysis using FUMA. Significant associations were identified for tissues relevant to male reproductive function. For “Malignant neoplasm of prostate”, associated genes were significantly enriched for expression in the prostate (P = 2.86 × 10^-6^) and testis (P = 2.60 × 10^-4^), and for “Hyperplasia of prostate” for gene expression in the prostate (P = 6.34 × 10^-3^), consistent with the tissue origin and endocrine involvement of the diseases.

Additional enrichment for “Malignant neoplasm of prostate” was noted in stomach (P = 3.83 × 10^-1^) and *esophagus mucosa* (P = 2.09 × 10^-4^), which reflects a similar gene expression profile between these tissues (31). Surprisingly, “Premature ejaculation” was linked to expression in the fallopian tube (P = 4.15 × 10^-4^), although a female reproductive tissue, it may indicate shared gene expression patterns with male reproductive tissues or involvement in broader reproductive signalling pathways.

### Genetic correlation

#### Pairwise genetic correlations

To investigate shared etiological architecture across male reproductive and genitourinary phenotypes, we conducted genetic correlation analyses using 29 phenotypes encompassing prostate inflammation, testicular and penile disorders, sexual dysfunction, infertility, and lifestyle traits such as BMI, smoking and alcohol use (Supplementary table 15).

At the genetic level, 28 pairwise correlations remained significant after FDR correction (P_FDR_ < 0.05; Figure 5). Hyperplasia of prostate exhibited strong positive genetic correlations with prostate-related conditions, including “Inflammatory disorders of prostate” (_rg_ = 0.86, P_FDR_ = 1.02 × 10^-24^), “Malignant neoplasm of prostate” (_rg_ = 0.34, P_FDR_ = 2.39 × 10^-9^), and “Prostate-specific antigen levels” (_rg_ = 0.57, P_FDR_ = 0.003). It also showed positive correlations with “Hydrocele”, “Spermatocele”, “Orchitis and epididymitis” and “Erectile dysfunction”. Similarly, “Erectile dysfunction” was genetically correlated with “Chronic prostatitis” (_rg_ = 0.61, P_FDR_ = 1.90 × 10^-5^), “Hyperplasia of prostate” (_rg_ = 0.31, P_FDR_ = 7.08 × 10^-5^), “Malignant neoplasm of prostate” (_rg_ = 0.30, P_FDR_ = 7.21 × 10^-4^), “Sexual dysfunction” (_rg_ = 0.95, P_FDR_ = 3.85 × 10^-11^), and “Testicular dysfunction” (_rg_ = 0.85, P_FDR_ = 1.21 × 10 ^-3^)].

**Figure 5.**
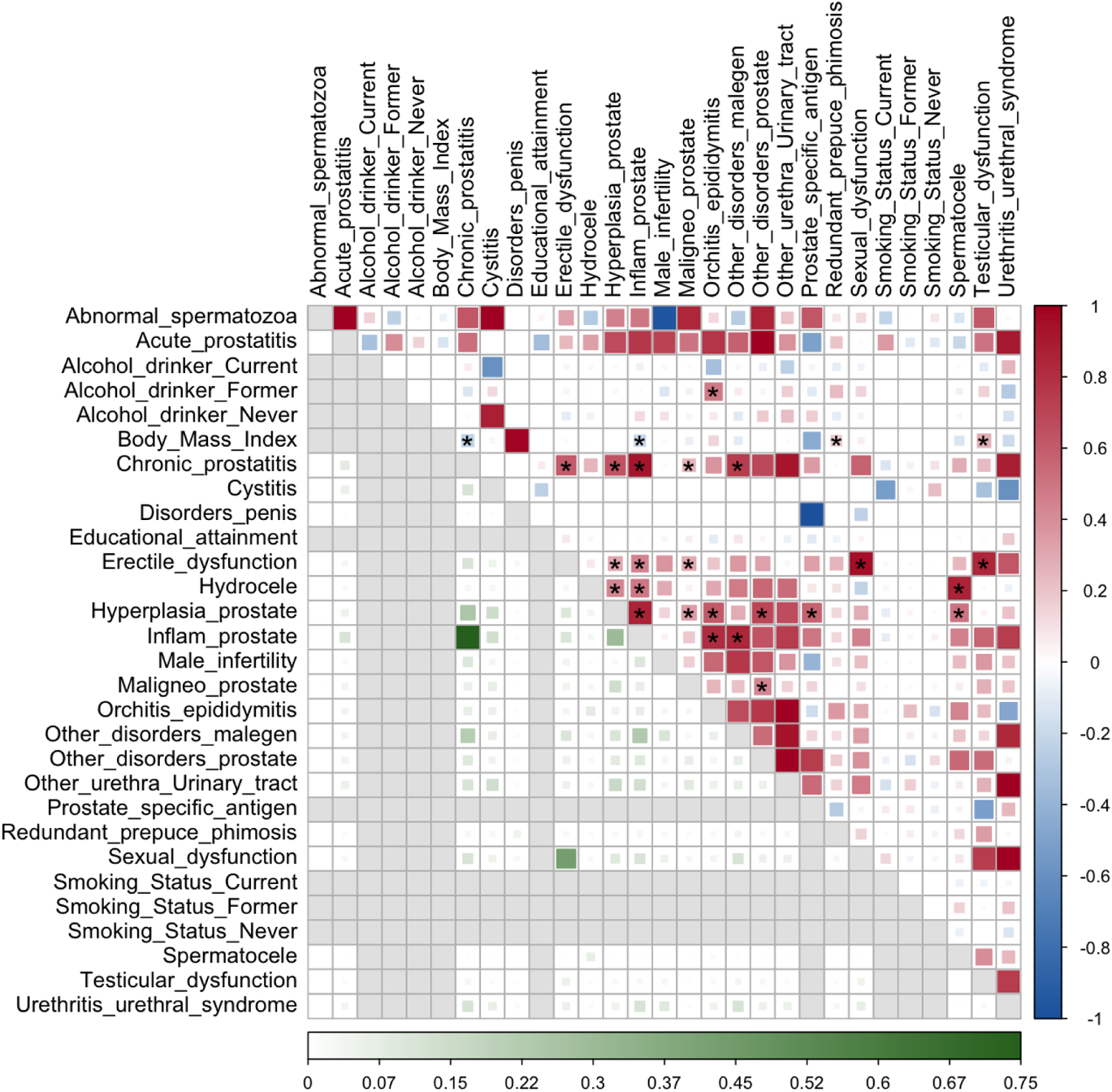
Results of Phenotypic overlap (bottom plot) and Genetic Correlation (upper plot) Analyses. The figure displays results for male reproductive binary diagnoses/traits (effective sample size >5,000). For the phenotypic overlap plot, darker green colours indicate a higher similarity/case overlap of the two phenotypes. Grey colour means that the phenotypic overlap was not computed for the phenotype pair. For the genetic correlation plot, the red colour indicates positive correlation, and blue indicates negative correlation. Correlation with FDR-adjusted P-values < 0.05 are denoted by asterisks in the upper plot. The size of the square corresponds to the respective estimated values. Inflam= Inflammatory disorders; Maligneo= Malignant neoplasm.

BMI showed significant negative genetic correlations with “Chronic prostatitis” (_rg_ = −0.23, P_FDR_ =0.0015) and “Inflammatory disorders of prostate” (_rg_ = −0.20, P_FDR_ = 0.0039), and positive genetic correlations with “Redundant prepuce, phimosis and paraphimosis” (_rg_ = 0.18, P_FDR_ = 0.00015), and “Testicular dysfunction” (_rg_ = 0.28, P_FDR_ = 0.025). Among other lifestyle traits, only “Alcohol drinker status: Former” was genetically correlated with “Orchitis and epididymitis” (_rg_ = 0.48, P_FDR_ = 0.03).

#### Reproductive hormones and other reproductive traits

In addition to the genetic correlation analyses based on the GWAS summary statistics generated in this study, we also performed analyses using publicly available summary statistics from the GWAS Catalog and from studies with larger sample sizes for reproductive traits and hormone levels (Supplementary table 16). This expanded approach revealed significant genetic correlations (FDR-adjusted P < 0.05) for 6 reproductive traits that had not reached statistical significance in our original analysis, as well as for 14 reproductive hormones (Figure 6; Supplementary table 17).

**Figure 6.**
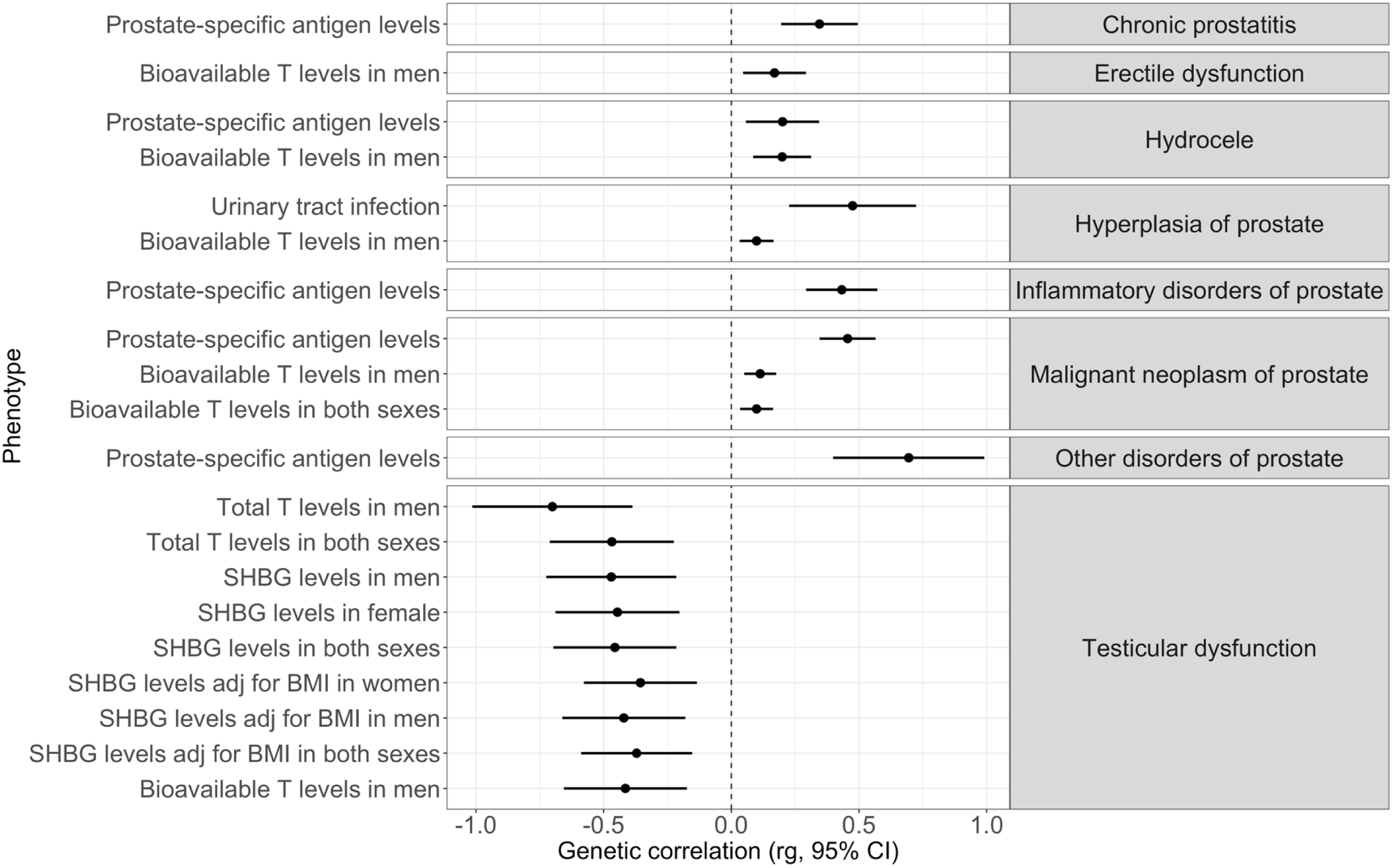
Genetic correlation for the male phenotypes and GWASs summary statistics for reproductive hormones/traits performed in other studies. The source of the GWASs can be found in the Supplementary table 17. Only genetic correlations that were statistically significant after FDR correction and were not reported in the pairwise genetic correlations between our GWASs are displayed in the figure. SHBG = sex-hormone binding globulin; T = testosterone; BMI = body mass index.

Among the newly identified associations for reproductive phenotypes, the most notable were those involving “PSA levels”, which showed strong positive genetic correlations with several prostate-related conditions, including “Malignant neoplasm of prostate”, “Inflammatory disorders of prostate”, “Other disorders of prostate”, and “Chronic prostatitis” (_rg_ = 0.34 - 0.69). “PSA levels” were also positively correlated with “Hydrocele” (_rg_ = 0.20, P_FDR_ = 4.79 × 10^-2^). In addition, urinary tract infection exhibited a positive genetic correlation with “Hyperplasia of prostate” (_rg_ = 0.47, P_FDR_ = 1.06 × 10^-3^).

Regarding reproductive hormone correlations, testicular dysfunction showed eight significant negative genetic correlations with total testosterone, bioavailable testosterone, and SHBG levels (_rg_ range: −0.47 to −0.36). Bioavailable testosterone levels in men were positively correlated with “Hydrocele” (_rg_ = 0.20, P_FDR_ = 8.70 × 10^-3^), “Hyperplasia of prostate” (_rg_ = 0.10, P_FDR_ = 2.10 × 10^-2^), “Malignant neoplasm of prostate” (_rg_ = 0.11, P_FDR_ = 5.29 × 10^-3^), and “Erectile dysfunction” (_rg_ = 0.20, P_FDR_ = 4.79 × 10^-2^).

#### Other phenotypes

Finally, we performed LDSC to estimate genetic correlations between the male reproductive phenotypes and traits from the CTG database. After FDR correction, seven phenotypes showed statistically significant genetic correlations: “Chronic prostatitis”, “Hydrocele”, “Hyperplasia of prostate”, “Inflammatory disorders of prostate”, “Malignant neoplasm of prostate”, “Orchitis and epididymitis” and “Redundant prepuce, phimosis and paraphimosis” (Supplementary table 18). The CTG traits were grouped into 4 categories: mental health, obesity/metabolic, reproductive, and other.

The mental health category included traits related to emotional symptoms (e.g. anxiety, nervousness, excessive worrying) and psychiatric diagnoses (e.g., depression, attention-deficit/hyperactivity disorder (ADHD), schizophrenia). Four male phenotypes (“Chronic prostatitis”, “Hyperplasia of prostate”, “Inflammatory disorders of prostate” and “Redundant prepuce, phimosis and paraphimosis”) exhibited genetic correlations with this category and all the genetic correlations were positive (ranging from _rg_ = 0.11 to 0.72).

In the obesity/metabolic category, genetic correlations with “Chronic prostatitis” and “Inflammatory disorders of prostate” were predominantly negative. In contrast, “Orchitis and epididymitis” and “Redundant prepuce, phimosis and paraphimosis” were mostly positively genetically correlated with traits in this category, except for “HDL cholesterol” and “Started insulin within one year diagnosis of diabetes” that showed negative genetic correlation with “Redundant prepuce, phimosis and paraphimosis”.

In the reproductive category, all 7 male phenotypes demonstrated positive genetic correlations. This category was mostly composed of diagnoses related to prostate, female reproductive system, and genitourinary tract. Most of the correlations in this analysis confirmed our previous findings from the pairwise genetic correlation analysis (Supplementary table 18). In addition to these replicated results, we identified novel genetic correlations, including between “Chronic prostatitis” and “Age at first birth” (defined in women) (_rg_ = 0.26, P_FDR_ = 8.47 × 10^-3^); “Female genital prolapse” and both “Hyperplasia of prostate” (_rg_ = 0.19, P_FDR_ = 4.17 × 10^-2^) and “Inflammatory disorders of prostate” (_rg_ = 0.35, P_FDR_ = 2.72 × 10^-2^); and “Abdominal and pelvic pain” with both “Hyperplasia of prostate” (_rg_ = 0.19, P_FDR_ = 1.34 × 10^-2^) and “Inflammatory disorders of prostate” (_rg_ = 0.60, P_FDR_ = 4.96 × 10^-5^).

### Phenotype-level correlation

Case-level co-occurrence was accessed across 19 phenotypes using the Jaccard Index and between male infertility and malignant neoplasms (Supplementary table 19). We found that male reproductive and urogenital disorders displayed varying degrees of case overlap, with Jaccard indices ranging from 0.0009 to 0.75. The highest overlap was found between inflammation-related conditions, such as “Chronic prostatitis” and “Inflammatory disorders of prostate” (Jaccard = 0.75), followed by “Erectile dysfunction” and “Sexual dysfunction” (Jaccard = 0.43). Moderate overlap was found between “Hyperplasia of prostate” and “Inflammatory disorders of prostate” (Jaccard = 0.30), and between “Chronic prostatitis” and “Hyperplasia of prostate” (Jaccard = 0.25). “Male infertility” was associated with increased odds of “Malignant neoplasms” (OR = 1.60, P = 4.47 × 10^-6^), indicating that infertile men had nearly twice the odds of having a cancer diagnosis compared with those without infertility.

## DISCUSSION

This large-scale genome-wide study of 64 male reproductive phenotypes was designed to advance understanding of the genetic architecture underlying a broad spectrum of genitourinary health in men. Genetic studies are essential for identifying biological mechanisms that cannot be captured through clinical or environmental data alone, enabling deeper comprehensions into disease etiology and opportunities for early risk stratification. We report 143 genome-wide significant lead variants, including 47 novel trait-specific associations and the first genome-wide insights into three phenotypes: “Abnormal spermatozoa”, “Redundant prepuce, phimosis and paraphimosis”, and “Testicular dysfunction”. Many of our associations replicate previously reported genetic findings, providing validation across multiple cohorts and supporting the robustness of these genetic signals. These findings have important implications for understanding the genetics of male reproductive health and disease and offer a foundation for future translational efforts in precision urology and andrology, a domain historically underrepresented in complex trait genetics.

By integrating multiple lines of evidence, we prioritised 328 candidate causal genes across the genomic risk loci, including several with known relevance to reproductive function, such as *CHEK2*, *KLK3*, *RSPO2* and others newly implicated in prostate phenotypes (44–46). The recurrence of specific genes across distinct yet related conditions (e.g., *FGFR2*, *POU5F1B*, *TERT*) highlights potential pleiotropic mechanisms in prostate development, function, and disease (47–49). Notably, *RSPO2* and *CHEK2*, both implicated in oncogenic and DNA repair pathways, respectively, may serve as functional links between reproductive function and cancer susceptibility (39,50).

Extending these gene-level insights, our analyses demonstrate the value of genetically bottlenecked populations for identifying population-enriched risk alleles that may be rare or absent in broader European cohorts. A prominent example is the missense variant rs200737258 at the *AR* locus, identified as novel genome-wide association signal for prostate cancer and markedly enriched in Finnish and Estonian populations. Given the central role of *AR* in prostate development and androgen insensitivity syndromes (51–53), this finding underscores the biological relevance of population-specific discoveries. More broadly, the enrichment of multiple lead variants in Finland and Estonia is consistent with the founder effects and reduced admixture increasing the frequency of otherwise rare alleles, thereby enhancing power for discovery (4,42). Similar patterns have been reported in FinnGen and the Estonian Biobank, where population-specific allele frequency structures have enabled the detection of clinically relevant variants not captured in pan-European analyses (10). Together, these results highlight the importance of including diverse European populations to improve variant discovery and genetic representation in large-scale human genetic studies.

Among the newly associated phenotypes, we identified distinct signals that broaden our understanding of the genetic architecture of male reproductive health. Notably, rs17632542 in *KLK3* (kallikrein related peptidase 3), which encodes PSA, was associated with “Abnormal spermatozoa”. This variant has known links with prostate cancer, PSA levels, and chronic prostatitis (9,54), and may affect sperm quality via its role in semen liquefaction (55,56). These findings suggest that variation in *KLK3* may contribute to both oncogenic and reproductive pathways through a shared biochemical mechanism.

The variant rs11198372 was significantly associated with “Redundant prepuce, phimosis, and paraphimosis”, and our gene prioritisation approach identified three genes (*EMX2*, *KCNK18*, and *ENO4*) with known reproductive phenotypes in knockout mouse models, suggesting plausible roles in human preputial development. *EMX2* knockout mice exhibit agenesis of internal male genitalia, including absence of the vas deferens, epididymis, and seminal vesicles, as well as agonadism in both sexes (57). *ENO4* deficiency leads to severe sperm flagellar defects and male infertility (58,59), while *KCNK18* mutants show abnormal and enlarged epididymis morphology. While none of these genes were prioritised based on coding or regulatory annotations at this locus, their shared reproductive phenotypes in mouse models support a potential developmental mechanism linking them to the studied trait. Functional validation will be essential to clarify whether these genes contribute to penile and foreskin morphology in humans.

A particularly noteworthy finding in our study is the potential interplay between immune-related genetic factors and metabolic traits influencing these conditions. Specifically, we identified a genome-wide significant association at the HLA region (rs12662632) and observed a positive genetic correlation with BMI, as well as with several other obesity/metabolic traits. The HLA region plays a pivotal role in immune system regulation and, especially the HLA Class II, has been implicated in various inflammatory and autoimmune conditions (60). In the context of phimosis, that is a chronic inflammation of the foreskin, it may be influenced by HLA-mediated immune responses, contributing to the development of fibrotic changes and foreskin constriction (61). Obesity is characterized by chronic low-grade inflammation, which can exacerbate immune responses and tissue remodelling processes (62). The positive genetic correlation between the traits suggests that higher adiposity may enhance susceptibility to foreskin pathologies through inflammatory pathways. Furthermore, studies have shown that obesity-related metabolic disturbances, such as diabetes mellitus, can impair skin integrity and healing, potentially influencing the progression of phimosis (63). This integrative understanding highlights the importance of considering both immunogenetic and metabolic factors in the etiology of foreskin pathologies.

Unexpectedly, we observed statistically significant negative genetic correlation between BMI and obesity/metabolic traits and both inflammatory diseases of prostate and chronic prostatitis. This is surprising given that prior studies have reported positive associations between obesity and prostatic inflammation (64). Obesity has been linked to systemic inflammatory states and alterations in local immune responses, which may influence prostate tissue remodelling and inflammatory profiles (65). However, an observational study reported an inverse association between BMI and prostatitis-like symptoms, although the authors attributed this finding to potential confounding by physical activity and did not pursue it further (66). While inconclusive, this evidence aligns with the direction of our genetic findings, which are less susceptible to behavioural confounding. These results suggest a more complex relationship between metabolic traits and prostate inflammation, potentially involving distinct mechanisms of genetic predisposition and environmental exposure. Further investigation is needed to clarify how obesity-related genetic factors might modulate prostate immune responses and inflammatory disease risk.

Beyond correlations among male phenotypes, we also observed positive genetic correlations between inflammatory prostate phenotypes and female traits related to reproductive timing and pelvic health. These associations may reflect shared genetic influences on biological aging and chronic inflammation, as leukocyte telomere length, a heritable marker of cellular aging, has been linked to systemic inflammation and age-related disease risk (67). In this context, age-dependent female traits such as genital prolapse and age at first birth, together with inflammatory prostate disorders in men, may partly capture common genetic susceptibility to aging-related inflammatory and connective tissue pathways rather than sex-specific pathology alone.

Consistent with previous evidence linking male infertility and cancer risk (36), we also observed a statistically significant positive association between Male infertility and malignant neoplasms, with infertile men showing 1.6-fold higher odds of cancer after accounting for age and population structure. This finding supports the notion of shared genetic or biological mechanisms predisposing to both impaired spermatogenesis and tumorigenesis.

Our findings reveal consistent genetic and phenotypic patterns across male reproductive and urological phenotypes, highlighting a shared genetic architecture among conditions traditionally viewed as distinct. In particular, the clustering of “Hyperplasia of prostate”, “Chronic prostatitis”, and “Erectile dysfunction” mirrors prior reports that link prostatic and sexual disorders through inflammatory and endocrine pathways, with chronic inflammation playing a central role in prostate disease development and progression (68).

“Erectile dysfunction” also correlated with broader sexual and structural disorders, suggesting common etiologies, potentially involving androgen or vascular dysfunction. The observed association between higher BMI and both “Testicular dysfunction” and “Redundant prepuce, phimosis and paraphimosis” aligns with prior work linking obesity to male hypogonadism and compromised reproductive health (69). Similarly, the genetic correlation between former alcohol use and epididymal inflammation may reflect persistent effects of alcohol-induced mitochondrial dysfunction, even after cessation (70).

Although we replicated many previous findings, this study has some limitations. First, since we do not have supporting data for the “Premature ejaculation” GWAS using only EstBB data, the results should be interpreted with care. Second, “Prostate-specific antigen levels” from EstBB should also be interpreted with caution, as they were extracted from electronic health records and likely reflect measurements taken in the context of clinical suspicion, which may influence their levels. Third, the definition of the phenotypes was solely based on ICD-10 and LOINC codes which may not capture the clinical and biological complexity of certain conditions, potentially masking heterogeneity or overlapping etiologies across phenotypes. Finally, this study was based on European ancestry cohorts, studies using other ancestries need to be done to confirm the association of these variants for the respective phenotypes in non-European populations. Together, our integrative analysis underscores the value of combining genetic and clinical data to disentangle the complex architecture of male reproductive health.

## CONCLUSIONS

This study presents the most comprehensive genome-wide analysis of male reproductive health to date, spanning 64 phenotypes and integrating data from over 500,000 individuals across EstBB, FinnGen, and UK Biobank. We identified 143 genome-wide significant variants, including 47 novel trait-specific associations and the first genetic signals for conditions such as “Abnormal spermatozoa”, “Redundant prepuce, phimosis and paraphimosis” and “Testicular dysfunction”. Through rigorous gene prioritisation and integrative analyses, including tissue-specific expression, functional annotation, and genetic correlation, we highlighted shared biological mechanisms underlying prostate, testicular, penile, and hormonal traits.

Our findings reveal extensive pleiotropy among male reproductive conditions, underscore the role of population-specific variants in discovery, and provide strong evidence for immune, endocrine, and developmental pathways contributing to genitourinary disease. The identification of novel loci in genes such as *CHEK2*, *RSPO2*, *KLK3*, *SHBG*, and *EMX2* further expands the catalogue of candidates for functional follow-up and potential clinical translation.

By addressing a critical knowledge gap in men’s health, this work lays a foundation for improved diagnostics, risk prediction, and therapeutic targeting in male reproductive disorders. Future efforts should prioritize longitudinal clinical integration, functional validation of candidate genes, and exploration of gene-environment interactions to advance precision medicine approaches in andrology and urology.

## ACKNOWLEDGEMENTS

This work was supported by the Estonian Research Council grants PRG1911, PRG1844, and PSG776; by the Ministry of Education and Research Centres of Excellence grant TK214 (Centre of Excellence in Genomics and Translational Medicine); and by the Estonian Ministry of Education and Research (project TEM-TA72). This project has received funding from the European Union’s Horizon Europe research and innovation programme under grant agreement No 101060011 and Project No. 2021-2027.1.01.24-0444. Views and opinions expressed are however those of the author(s) only and do not necessarily reflect those of the European Union or European Research Executive Agency. Neither the European Union nor the granting authority can be held responsible for them.

The research was conducted using the Estonian Center of Genomics/Roadmap II funded by the Estonian Research Council (project number TT17). The activities of the EstBB are regulated by the Human Genes Research Act, which was adopted in 2000 specifically for the operations of the EstBB. Individual level data analysis in the EstBB was carried out under ethical approval 1.1-12/624 from the Estonian Committee on Bioethics and Human Research (Estonian Ministry of Social Affairs), using data according to release application 6-7/GI/29022 from the Estonian Biobank. Data analysis was carried out in part in the High-Performance Computing Center of University of Tartu. We want to acknowledge the participants and investigators of the EstBB, FinnGen, and UKBB studies.

The GTEX data used for the analyses described in this manuscript were obtained from eQTL Catalog on October 1, 2024. The Genotype Tissue Expression (GTEx) Project was supported by the Common Fund of the Office of the Director of the National Institutes of Health, and by NCI, NHGRI, NHLBI, NIDA, NIMH, and NINDS.

## DATA AVAILABILITY

Summary statistics from UK Biobank and FinnGen used in this study are accessible through the UKBB PheWeb portal (https://pheweb.org/UKB-TOPMed/phenotypes) and FinnGen PheWeb (https://r7.finngen.fi/ and https://r8.finngen.fi/), respectively. Full GWAS summary statistics and meta-analysis will be deposited in the GWAS Catalog upon acceptance. Genome-wide association analyses were conducted using REGENIE (version 3.2), and meta-analyses were performed with GWAMA. Additional resources consulted include the Mouse Genome Database (http://www.informatics.jax.org), GTEx Portal (https://gtexportal.org/home/), eQTL Catalog (https://www.ebi.ac.uk/eqtl/), GWAS Catalog (https://www.ebi.ac.uk/gwas/), and the Complex Trait Genetics Virtual Lab (https://vl.genoma.io/).

## Notes

### Competing Interest Statement

The authors have declared no competing interest.

### Summary of Updates

Revision submitted to correct title capitalisation.

