## Supplementary Figure 1 for "Genome-wide analysis of 64 male reproductive phenotypes reveals novel loci and shared biological pathways"

**Supplementary Figure 1.** Variants showing high heterogeneity (I^2^>70%). Numbers indicate the effect allele frequence (EAF) in each cohort.

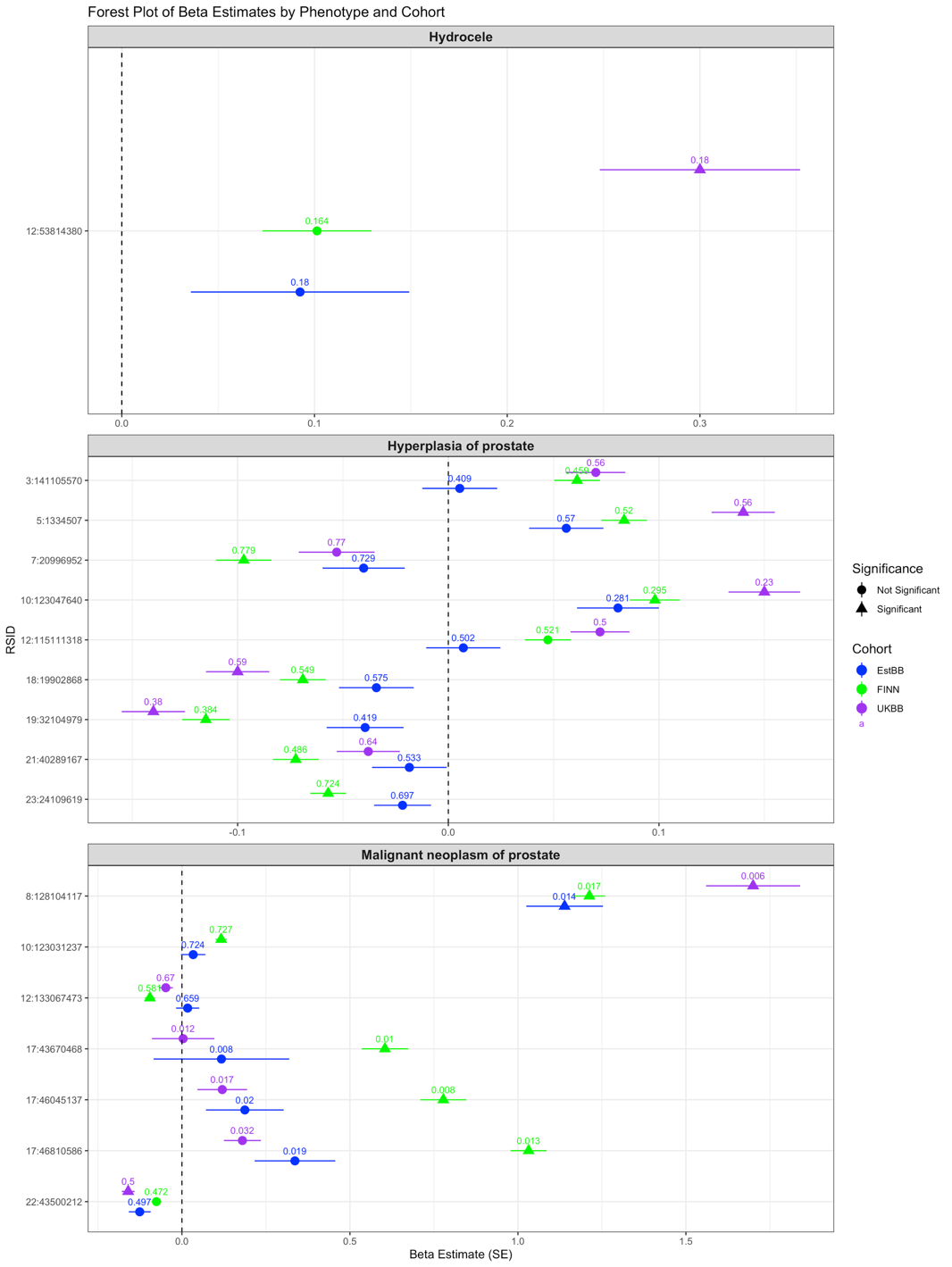
